# COVID-19 control measure effects suggest excess winter mortality is more sensitive to infection control than warmer temperatures

**DOI:** 10.1101/2020.12.19.20248531

**Authors:** Lucy Telfar-Barnard, Michael G Baker, Amanda Kvalsvig, Nick Wilson

**Affiliations:** Co-Search COVID-19 Research Collaborative, Department of Public Health, University of Otago, Wellington, New Zealand; BODE^3^ Programme, Department of Public Health, University of Otago, Wellington, New Zealand

## Abstract

**Background:** Excess winter mortality (EWM) has been attributed to both seasonal cold exposure, and to infectious disease. In 2020, New Zealand’s border management and lockdown measures successfully eliminated community transmission of SARS-CoV-2, and also largely eliminated influenza and many other respiratory viruses. This study investigates the contribution of infections and temperature to EWM and typical extended winter (May to October) deaths in this natural experiment created by New Zealand’s COVID-19 pandemic response.

**Methods:** We used age-standardised weekly deaths to measure EWM 2011 to 2019, then used historical patterns to estimate high, medium and low scenario 2020 EWMs. We then modelled typical year and 2020 heating degree day: mortality relationships to estimate relative contributions of cold temperature and infection to typical EWEDs.

**Results:** EWM 2011 to 2019 averaged 14.7% (low 11.4%, high 20.9%). In contrast, 2020 EWM was estimated at 1.6%, 2.7%, or 3.8% under high, medium, and low spring-summer mortality scenarios. Between 2011 and 2019, temperature was estimated to explain 47% of extended winter deaths, and infection 27%, with the remaining 26% attributable to the interaction between infection and temperature.

**Discussion:** The society-wide response to COVID-19 in 2020 resulted in a major reduction of winter mortality in this high-income nation with a temperate climate. In addition to influenza, other respiratory pathogens likely also make a significant contribution to EWM. Low cost protection measures such as mask wearing (eg, in residential care facilities), discouragement of sick presenteeism, and increased influenza vaccine coverage, all have potential to reduce future winter mortality.

**Research in context:** *Evidence before this study:* Excess winter mortality (EWM) is a widely observed phenomenon, commonly attributed to physiological responses to short and long-term outdoor and indoor cold exposure (and associated increased air pollution); other seasonal physiology changes; and higher incidence of some infectious diseases. Previous estimates of EWM in New Zealand range from 10.3% to 25.6%, with influenza estimated to make up roughly a third of that excess. Internationally, deaths attributable to cold temperatures are also found outside the traditional winter period, with influenza making a large contribution to cold temperature deaths.

*Added value of this study:* This study finds that following a successful COVID-19 elimination strategy, which simultaneously prevented the annual winter influenza season, and likely other winter respiratory infections, New Zealand is likely in 2020 to experience less than a third of the usual winter mortality excess. Further, this study for the first time estimates the relative contributions of cold temperature and infection, and the interaction between the two, to New Zealand winter deaths. We estimate that of the 9.5% fewer deaths than in typical years recorded between 1 May and 31 October 2020, 92.5% were prevented by infection control measures; 1.4% by the 1.14°C warmer than average winter; and 6.1% by the interaction between infection and low temperature.

*Implications of all the available evidence:* Influenza and other infectious respiratory pathogens appear to make a much larger contribution to winter mortality than previously recognised. Low cost protection measures such as mask wearing (eg, in residential care facilities), discouragement of sick presenteeism, and increased influenza vaccine coverage, all have potential to reduce future winter deaths, and lower overall annual mortality rates.

## Background

Excess winter mortality (EWM) describes the regular pattern of higher death rates in winter than non-winter. EWM has been observed worldwide for centuries, and has been variously attributed to physiological responses to seasonal cold exposure, whether outdoors or in poor quality housing; other seasonal physiological changes, such as reduction in vitamin D levels; and higher incidence of some infectious diseases.

EWM is commonly reported using Curwen’s index,^1^ which measures EWM as the percentage difference between mortality rates for the four winter months and the preceding and subsequent four month non-winter periods combined. EWM is easy to communicate and may capture seasonal effects missed in some temperature-mortality models.

However, EWM ratios can oversimplify or hide the true effect of winter seasonal effects on mortality.^2 3^ Associations between outdoor cold and increased mortality are now commonly measured using distributed non-linear lag models, measuring mortality increase per degree below a location- and/or cause-specific minimum mortality temperature;^4^ or per heating degree day (HDD).^3^ Such models routinely find cold-associated deaths occurring outside the three (or four) month winter period, with proportions of non-winter cold-associated deaths, and the degree to which EWM captures them, varying by country.

Influenza is broadly acknowledged to make up a large part of the annual excess winter death burden,^5^ though deaths may present as circulatory or other medical events in addition to pneumonia and respiratory death. However, circulatory deaths are also associated with cold temperature, making the respective contribution of infection and temperature to winter and broader health burdens difficult to disentangle.

In New Zealand, a temperate high-income island nation in the Southern Hemisphere, historical estimates of EWM have ranged from 10.3% to 25.6% (average 18.4%) for the period 1980 to 2000,^6^ and averaged 16.7% for the period 2000 to 2006.^7^ These estimates are consistent with levels in temperate European countries,^3^ as well as older estimates for Chile, which is at similar latitudes to New Zealand; and for Australia.^8^

A 2016 study of influenza and pneumonia mortality in the largest New Zealand city of Auckland, found a significant elevation in pneumonia and influenza deaths up to 19 days after cold and/or dry days.^9^ The cold temperature-to-mortality relationship has not otherwise been measured in New Zealand, possibly reflecting challenges securing appropriate data from a relatively small population spread over a climatically diverse country.

International estimates of seasonal influenza’s contribution to annual deaths range from 4.3 to 30.4 deaths per 100,000, with a pooled estimate of 13.9 (95%CI: 13.8 – 13.9). On average only 23 New Zealand deaths per year are coded as influenza, but modelled estimates of New Zealand’s influenza-attributable mortality rates range from 4.6 to 26.0, and averaging 13.5 (95%CI: 13.4 – 13.6) deaths per 100,000; a total of 498.8 (95%CI 496.6 – 501.0) deaths per year.^10^ However, the potential unattributed role of other infectious diseases in the annual winter or cold mortality burden does not appear to have been explored.

In 2020, New Zealand adopted an elimination strategy in response to the threat of the COVID-19 pandemic, involving a short, intense lockdown from March to May, combined with border management and other control measures (eg, testing, contact tracing etc).^11^ This approach successfully eliminated SARS-CoV-2 transmission in the community. It also largely eliminated seasonal influenza and many other respiratory viruses,^12^ and reduced overall mortality by 11%.^13^ Similar elimination of seasonal influenza from COVID-19 control strategies has been reported in Argentina, Australia, Chile, Paraguay and South Africa.^14^

In the months following control measure deployment, New Zealand also experienced its warmest winter on record, 1.14°C above average.^15^

In this study we aimed to estimate the effect of New Zealand’s COVID-19 response on both projected EWM for 2020, and on a broadly estimated temperature-mortality gradient, using the COVID-19 elimination strategy ‘natural experiment’, and the warmer winter, to assess the contribution of infections to excess winter mortality, and the relative contributions of infection and temperature to annual deaths.

## Methods

### Deaths

We used Statistics New Zealand (SNZ) weekly mortality data for the period 1 January 2011 to 28 November 2020.^16^ We excluded two sudden mass death events involving the 22 February 2011 Christchurch earthquake and the 15 March 2019 mosque shootings,^17^ to avoid underestimating the winter effect in those years. We age standardised weekly deaths to SNZ 2018 New Zealand population estimates, across four age groups (under 30 years, 30 to 59 years, 60 to 79 years, and 80 years and over), using a combination of population and mortality distributions to remove year-on-year trends in mortality rates. We then smoothed age-standardised weekly rates over days.

### EWM

As per Curwen’s index,^1^ smoothed daily deaths per day were allocated to winter (June to September in the Southern Hemisphere) and non-winter (February to May, October to January) months.

For the 2011 to 2019 period, we calculated average and smoothed maximum and minimum mortality rates per million people per day of year. We then used mortality rates to measure EWM for each complete year, and estimated the potential 2020 EWM under three scenarios. Actual mortality rates were used for 1 February to 28 November, and the following scenarios were used to estimate mortality rates from 29 November 2020 to 31 January 2021:

1. Rates decrease to meet the 1 January smoothed high, then track smoothed high.
2. Rates decrease to meet the 1 January average, then track average.
3. Rates decrease to meet the 1 January smoothed low, then track smoothed low.

### Heating degree days

We identified 28 geographically dispersed weather stations with hourly mean temperature data that were 100% complete for the study period in the New Zealand National Institute of Water and Atmospheric Research Cliflo National Climate Database. Using linear regression with smoothed daily mortality rates, we identified the Dargaville 2 Ews weather station (−35.93145 S, 173.85317 E) as showing the best linear association (ie, highest R^2^) between hourly minimum temperature and typical year mortality data (excluding 2017 as an abnormally high influenza year, and 2020 as the year of interest). Next, we used a heuristic approach to first identify how many days’ lag provided the closest association between temperature and mortality, and then to identify the thermal comfort threshold, using second degree polynomial regression with typical year mortality rates and daily sums of hourly HDDs (in 0.1 degree increments), to identify the point at which the lowest modelled mortality rate coincided with the lowest actual mortality rate.

We then modelled two HDD: mortality relationships. First, we modelled the typical year(1 February to 31 January) HDD: mortality association, with second degree polynomial regression providing the best fit. Daily mortality rates were modelled for aggregated typical years, and for 2020, providing an estimate of what 2020 daily mortality rates might have arisen over an extended Southern Hemisphere winter (1 May to 31 October) in normal circumstances with 2020 temperatures. Second, we modelled the relationship between 1 May to 29 November 2020 mortality rates and HDDs, with a linear regression providing the best fit, and used the model to estimate what mortality rates might have occurred with typical year temperatures had those years also had 2020 infection control measures. Finally, we compared the results of the two models, and the model constants, to estimate the relative contributions of infection and temperature to New Zealand’s annual extended winter deaths.

### Patient and public involvement statement

Patients and the public were not involved in the design, or conduct, or reporting, or dissemination plans of this research.

### Ethics approvals

Ethical approval for this study was granted by the University of Otago, approval number HD19/073.

### Findings

The average EWM for 2011 to 2019 in the New Zealand population was 14.7%, with a low of 11.4% in 2011 and a high of 20.9% in 2017. These represent an average of 1537 excess winter deaths per year (low 1219, high 2226).

Peak daily mortality ranged from 22.1 deaths per million people per day in 2013, to 25.5 in 2017 (Figure 2). Average, high and low EWMs by age group are shown in Table 1.

**Table 1.** Average, high and low EWM by age group, in the pre-pandemic years: 2011 to 2019.

| **Age group** | **Mean EWM** | **Low EWM (year)** | **High EWM (year)** |
| --- | --- | --- | --- |
| Under 30 years | 4.3% | -4.6% (2019) | 16.0% (2017) |
| 30 to 59 years | 4.9% | 0.2% (2018) | 9.8% (2012) |
| 60 to 79 years | 10.6% | 7.0% (2016) | 13.4% (2019) |
| 80+ years | 20.5% | 14.6% (2013) | 31.2% (2017) |

New Zealand daily mortality rates were within usual ranges prior to its first COVID-19 case in late February 2020, and remained unremarkable until near the end of a stringent 25 March to 13 May lockdown. Deaths then began to diverge noticeably from their usual pattern. While the 2020 peak of 19.2 deaths per million people per day fell within the winter period (the week to 19 July), this peak was well below all previous peaks. After the first nationwide lockdown, apart from one week in June and one in July, mortality rates remained consistently below usual minima until near the end of the study period (Figure 1).

**Figure 1.**
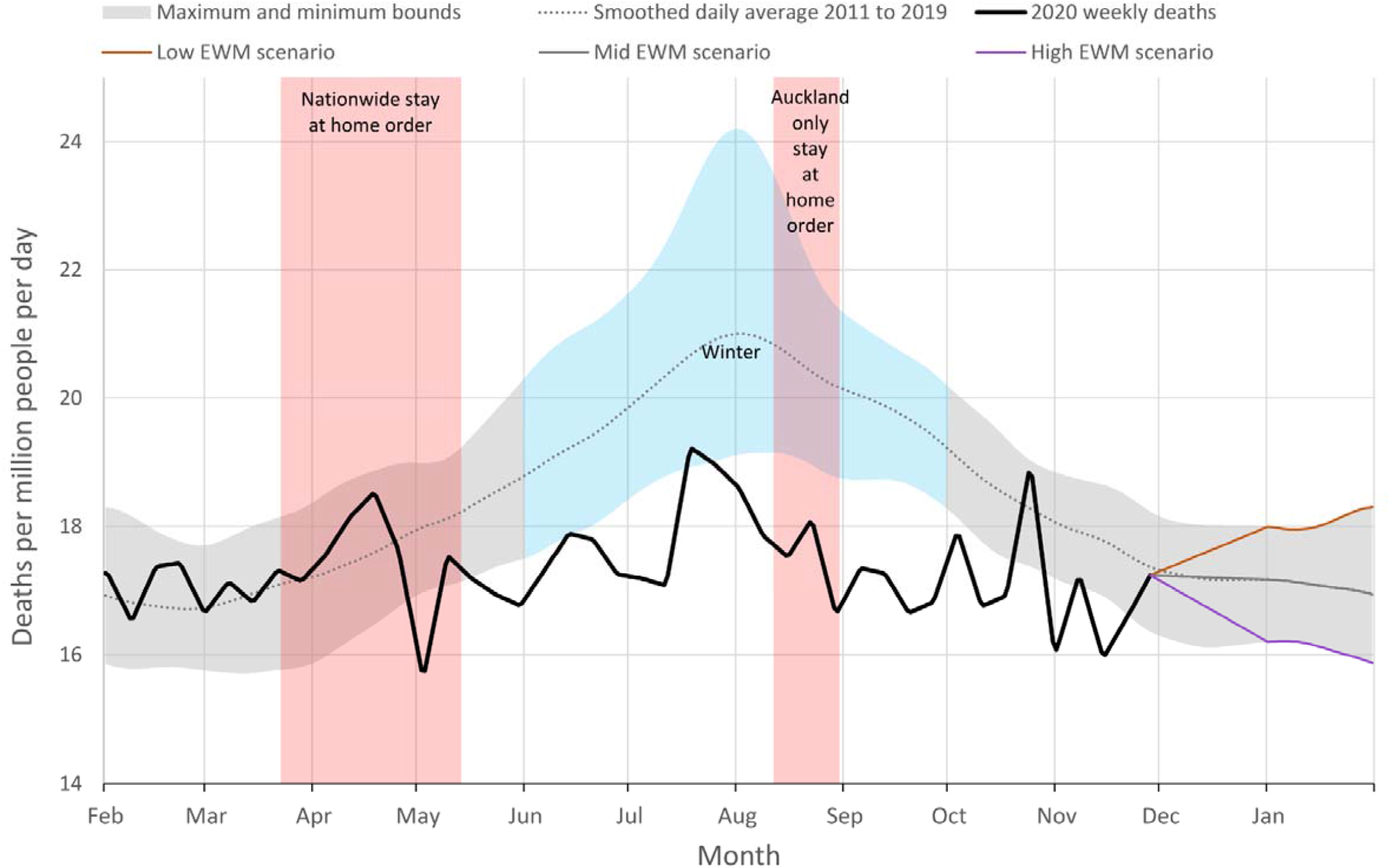
Age-standardised smoothed weekly deaths per million people per day in New Zealand, average and range, 2011 to 2019; observed 2020; and then estimated under high, medium and low EWM scenarios; February to January. (Note: Auckland is the largest New Zealand city with around a third of the NZ population).

**Figure 2.**
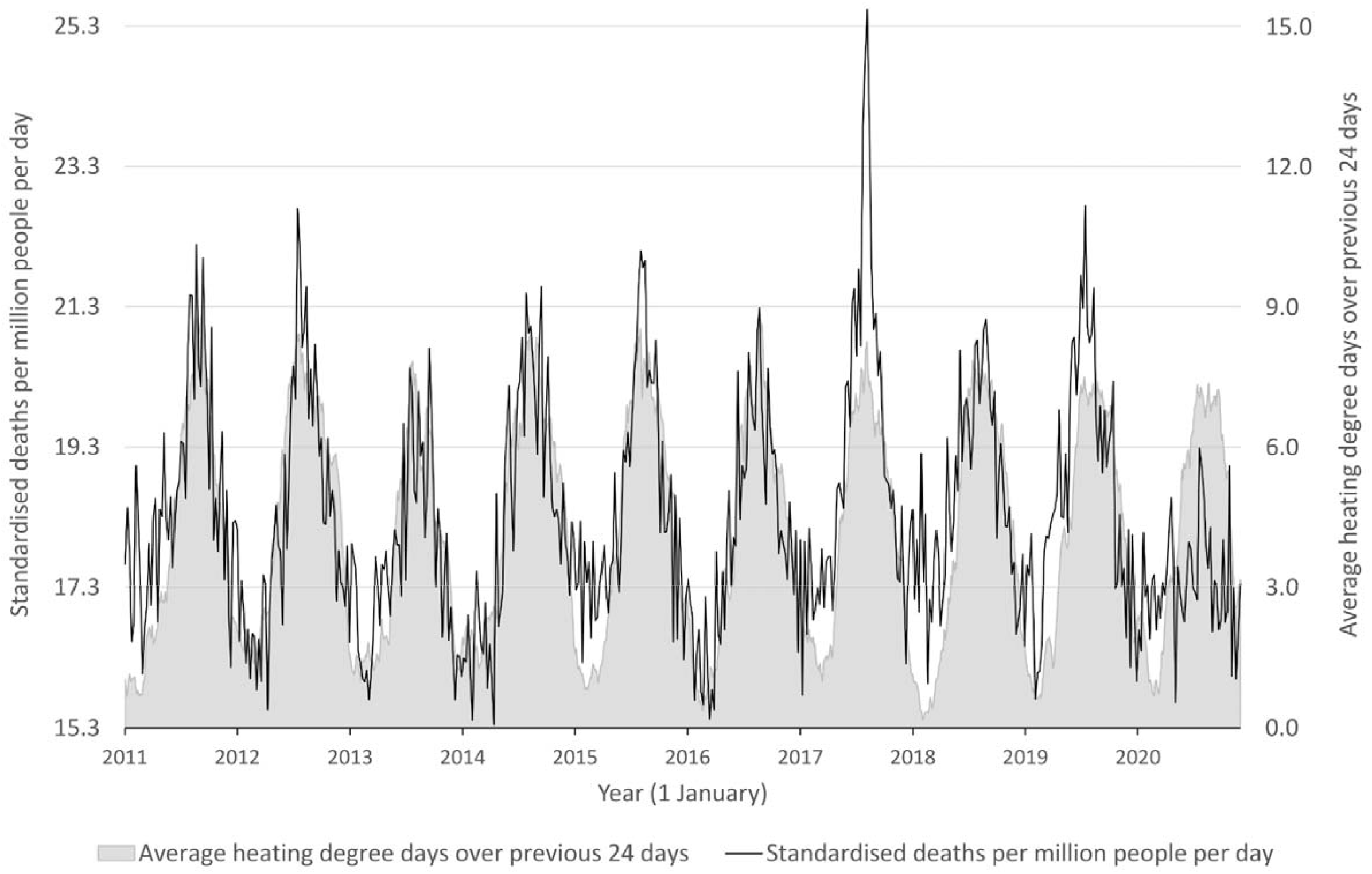
Average heating degree days and smoothed weekly deaths in New Zealand, 1 January 2011 to 22 November 2020.

Across the low (1), medium (2), and high (3) scenarios tested, the EWM for 2020 was 1.6%, 2.7%, or 3.8%. Expressed as absolute numbers, these represent 34; 56; or 79 excess winter deaths.

The Dargaville 2 Ews station is located in the northern (warmer) part of New Zealand. The annual average hourly mean temperature for the station 2011 to 2019 inclusive was 15.5 °C. For HDDs, a 24 day lag and a 19.4°C threshold were identified as providing the best fit across data for typical years.

The association between HDDs (h) and mortality rates per person per day (d) in a typical year was modelled to be: d = 0.059h^2^+0.010h+16.82 (R^2^=0.7293). In 2020 the relationship between deaths and temperature was linear rather than polynomial (d=0.256h+15.886), meaning an increase of 0.26 deaths per million people per day for each additional average HDD over the previous 24 days (95%CI: 0.20 – 0.31, p<0.001) (Figure 3). In a typical year, the equivalent figure for a linear fit was 0.53 deaths per million people per day (95%CI: 0.51 – 0.54, p<0.001).

**Figure 3.**
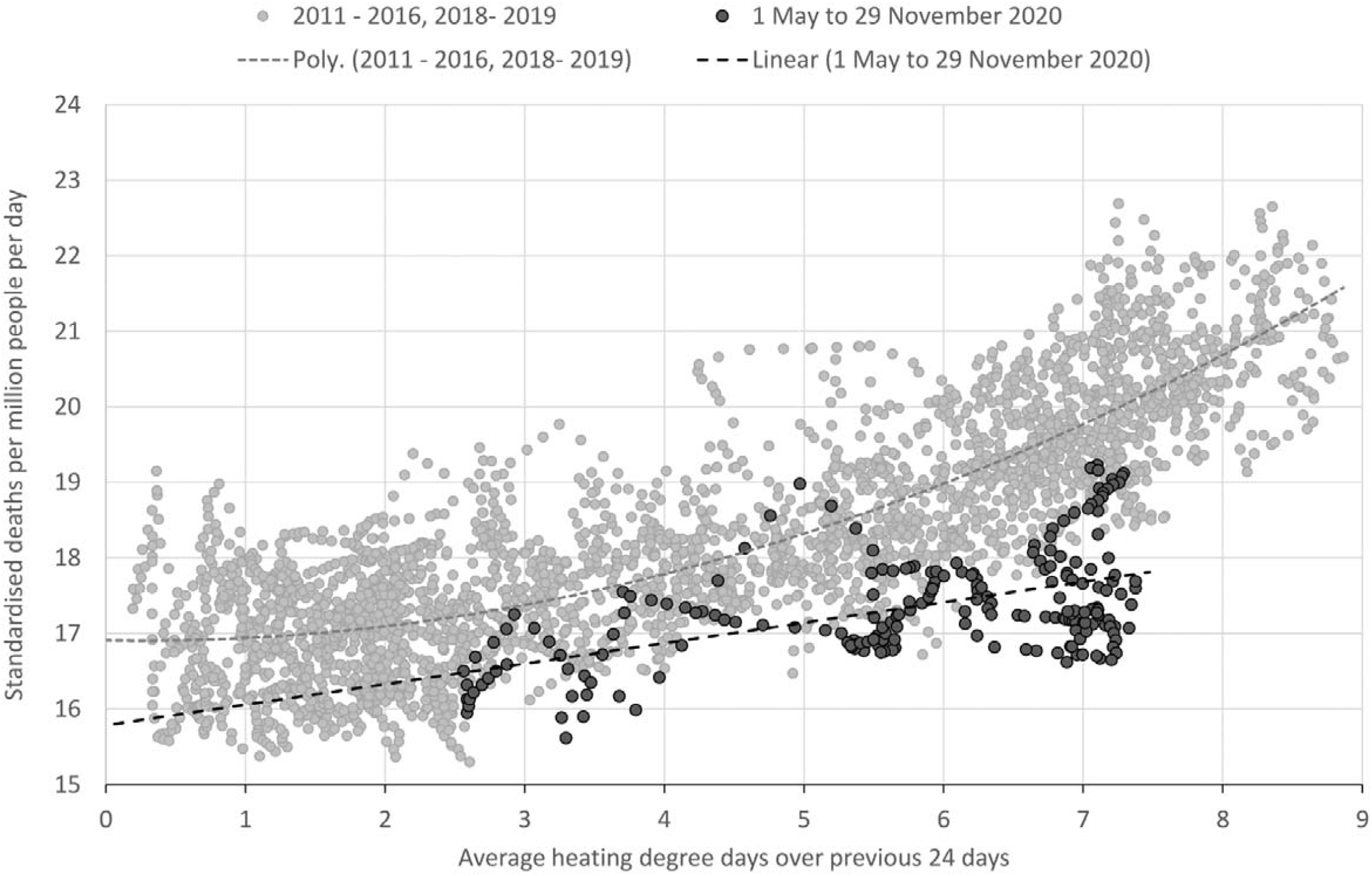
Deaths per million people per day and average heating degree days (HDD), and HDD to mortality relationships in New Zealand, typical years and 1 May – 22 November 2020. *Typical years = 1 February 2011 – 31 January 2017, 1 February 2018 – 31 January 2020.

Modelled differences in HDD-associated mortality rates and deaths between typical years and 2020 are shown in Table 2.

**Table 1.**
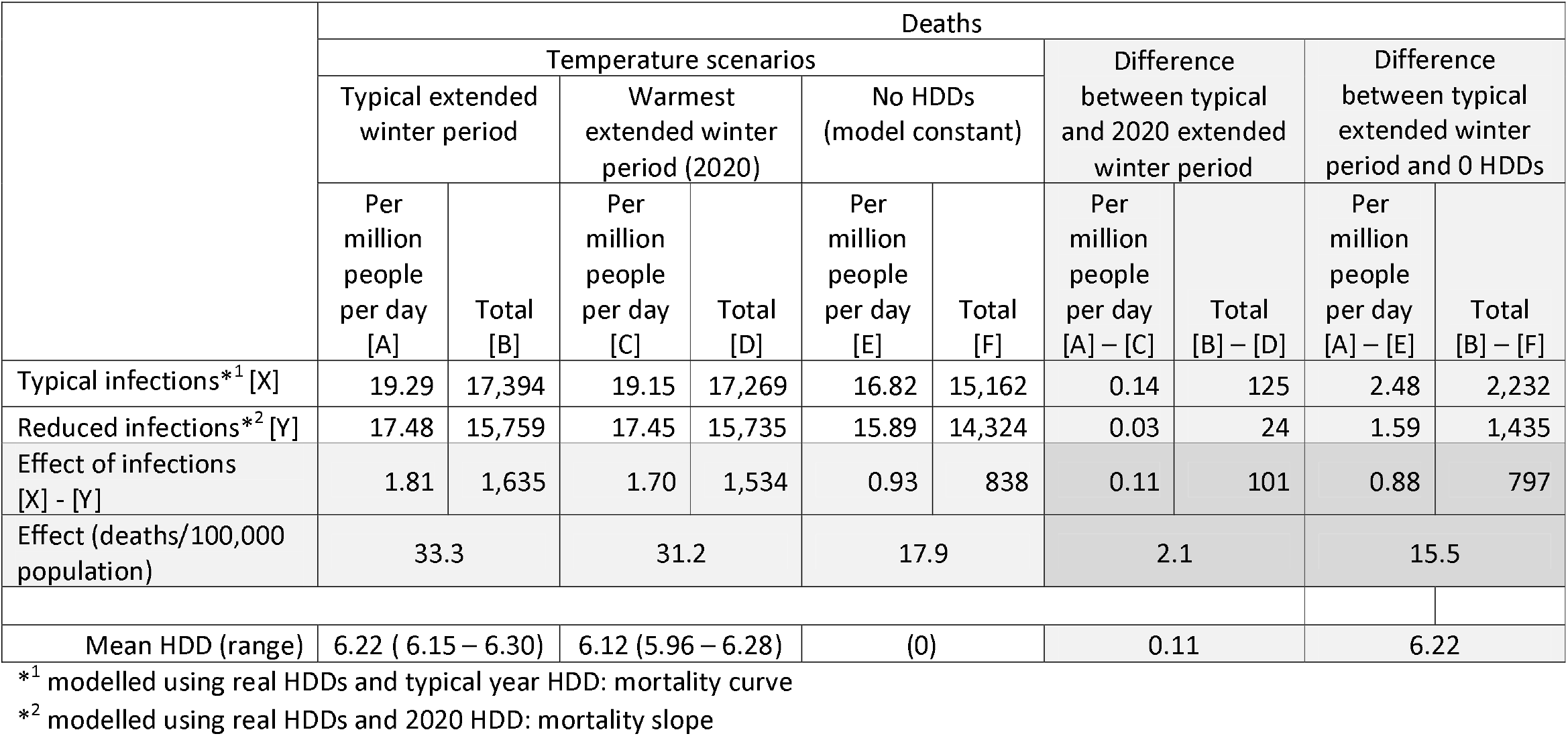
Relative contributions of HDDs, infection, and interactions, to annual extended winter deaths.

In a typical year, models estimated 19.29 deaths per million people per day (17,394 deaths) between 1 May and 31 October. Using the typical year model with 2020 temperatures over the same period, we modelled 19.15 deaths per million people per day (17,269 deaths). The constant for the typical year model was 16.82 deaths per million people per day (15,162 deaths).

Alternatively, using the 2020 HDD: mortality relationship with typical year temperatures, to estimate what previous years’ deaths might have been with similar infection prevention measures, we would have expected to see 17.48 deaths per million people per day (15,759 deaths) between 1 May and 31 October 2020, while equivalent figures for 2020 were 17.45 deaths per million people per day (15,735 deaths). The constant for the 2020 model was 15.89 deaths per million people per day (14,324 deaths).

For 1 May to 31 October 2020 mortality rates, we found 1,659 fewer deaths (1.84/million people/day) than in typical years, a reduction of 9.5%. We estimate 92.5% of those deaths (n=1,534; 1.70/million people/day) were prevented by infection control measures; 1.4% (n=24; 0.03/million people/day) by the warmer than average winter; and 6.1% (n=101; 0.11/million people/day) by the interaction between infection and temperature.

Combining 2020 results with estimates for zero HDD constants, we estimate a total of 3.40 cold- and infection-attributable deaths per million people per day (17.6% excess; 3,070 deaths) in an extended winter with typical temperatures and infection levels. Of these, 1.59 deaths per million people per day (47% of the excess, 1,435 deaths) might be attributed to cold temperature alone, and 0.93 deaths per million people per day (27%; 838 deaths) to infection, with an additional 0.88 deaths per million people per day (26%; 797 deaths) attributable to the interaction between infection and temperature. Modelled deaths per million people per day 1 February to 31 January, with and without infection control and/or cold temperatures are shown in Figure 4.

**Figure 4.**
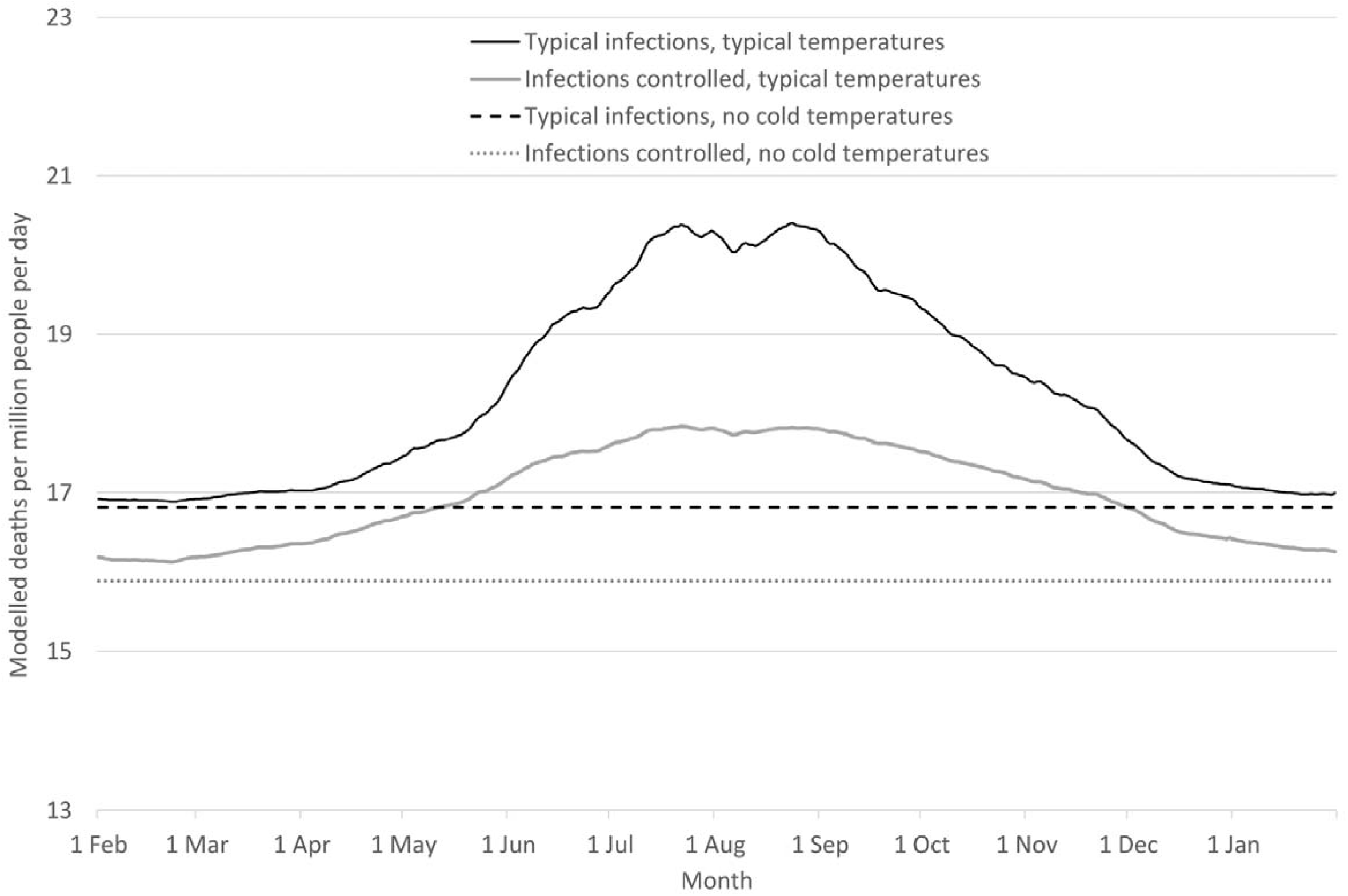
Modelled deaths per million people per day 1 February to 31 January, with and without infection control and/or cold temperatures.

### Interpretation

We observe that the identified HDD threshold of 19.4 °C was higher than the WHO recommended minimum indoor temperature of 18 °C, ^18^ a threshold commonly used in other heating degree day calculations.^3^ However, the Dargaville 2 Ews station, in the Northland region, is at the warmer end of New Zealand. While its annual average temperature of 15.5°C is lower than Auckland’s 2011 to 2018 average of 15.9°C, it is higher than New Zealand’s reference composite average temperature of 13.0 °C, ^19^ meaning that a 19.4 °C threshold in Dargaville would likely transpose to a lower threshold in many other parts of the country. As 96% of the population live in areas south of (and usually cooler than) Northland, Dargaville’s 19.4 °C threshold is not an indicator of likely thresholds in other parts of New Zealand.

The numbers of excess winter deaths in 2020 under all three EWM scenarios are markedly less than the average of 1537 in the preceding years. These estimates strongly suggest that New Zealand’s EWM for 2020 is historically low, due to winter deaths prevented rather than high summer deaths. With mortality rates continuing to run below usual levels in the most recent data, the final estimate for EWM for 2020 looks more likely to fall in the mid to high scenario range (2.7 – 3.8%), but even this range is less than a third of the usual 14.7% excess.

Meanwhile, the comparison of 1 May to 31 October temperature mortality relationships suggests that COVID-19 control measures and a warmer winter have reduced combined HDD- and infection-associated deaths by 1,659 deaths (1.84/million people/day), with 92.5% of those deaths (n=1,534; 1.70/million people/day) prevented by infection control measures; 1.4% (n=24; 0.03/million people/day) by the warmer than average winter; and 6.1% (n=101; 0.11/million people/day) by interaction between infection and temperature. Further, model constants suggest that even in a hypothetical period with no HDDs, 0.93 deaths per million people per day (838 deaths) may be attributable to infection, and a further 0.88 deaths per million people per day (797 deaths) attributable to temperature-infection interaction. These results also suggest that overall in an extended (six month) winter, 12.8% of deaths are attributable to cold temperature, and that 35.6% of those cold-attributable deaths could be prevented through fully effective prevention control.

However, while we have attributed the non-HDD-related reduction in mortality to reduced infection, other factors may also have contributed to reduced mortality. As Kung et al also note, mortality reductions following COVID-19 control measures may also have been due to “fewer deaths from road traffic accidents, occupational causes, air pollution, and postsurgical complications”. ^13^ We also observe that the New Zealand Government in 2020 doubled the amount of its Winter Energy Payment provided to low-income and superannuitant households, which with 2020’s warmest winter on record, could also have reduced the impacts of indoor cold exposure and poor housing. This payment, as well as the warmer winter, may have also reduced ambient air pollution from the burning of wood or coal for home heating.

There were also methodological limitations to this study: with a single temperature source generalised over a large climate range, and weekly deaths smoothed to produce daily estimates, the models cannot capture the full association between HDDs and deaths, such that the temperature-attributed deaths are likely to have been underestimated, and thus the infection-attributed deaths overestimated.

However, it is fair to hypothesise that a larger part of 2020’s EWM or extended winter health gains are attributable to prevention of respiratory infections than to the warmer winter. Influenza has previously been estimated to account for around 500 deaths a year,^20^ or roughly a third of the annual winter excess. Findings here suggest that other infectious pathogens, particularly those that cause death from pneumonia or precipitate fatal cardiovascular events, are also likely to make a large and currently unattributed contribution not only to winter deaths, but to cold-associated and baseline deaths outside the winter period. The longer the effects of New Zealand’s COVID-19 control measures continue in some form (eg, various levels of increased physical distancing and mask use on public transport), the greater will be our ability to assess that contribution.

Previous work has found EWM in New Zealand to be equally shared across ethnicities, but differentiated by age, housing tenure and, particularly for infectious disease, by income. ^21^ Overseas research has also found ethnic differences in cold-attributable deaths. ^22^ Future research may identify which groups benefited most from New Zealand’s COVID-19 response induced life gain, what categories of disease and injury contributed most, and how long the benefit persisted.

These findings support the need to continue efforts to improve thermal comfort and safety indoors and when going outdoors in winter, and other efforts to reduce cold exposure, to reduce cold-associated illness and death.

Meanwhile, although lockdowns are not feasible to address future endemic infection-related seasonal mortality, New Zealand’s 2020 experience suggests that lower-cost COVID-19 prevention measures deserve further consideration. These measures include encouraging mask-wearing in winter (eg, on public transport and when visiting aged residential care facilities) and strong employer policies discouraging sick presenteeism, as well as increased influenza vaccine coverage. Such policies could make further inroads into eliminating death’s annual winter harvest.

## Data Availability

Data used in the study have not been shared as all data used are publicly available. Mortality data used are available from the Statistics New Zealand COVID-19 Data Portal (https://www.stats.govt.nz/experimental/covid-19-data-portal). Temperature data used are available from the New Zealand National Institute of Water and Atmospheric Research CliFlo database (https://cliflo.niwa.co.nz/). Population data used are available from Statistics New Zealand (http://nzdotstat.stats.govt.nz).

## Contributors

LT-B collated the data, planned the analysis, prepared and analysed the data, and drafted and revised the paper. She is guarantor. MGB contributed to the conception of the study, and revised the paper. AK revised the paper. NW contributed to the conception of the study, contributed to the analytical plan, and revised the paper.

## Dissemination

This study used publicly available all-cause mortality data, meaning that dissemination to participants or patient organisations was not possible.

